# Tracking SARS-CoV-2 in rivers as a tool for epidemiological surveillance

**DOI:** 10.1101/2021.06.17.21259122

**Authors:** María Noel Maidana-Kulesza, Hugo Ramiro Poma, Diego Gastón Sanguino-Jorquera, Sarita Isabel Reyes, María del Milagro Said-Adamo, Martín Mainardi Remis, Dolores Gutiérrez-Cacciabue, Héctor Antonio Cristóbal, Mercedes Cecilia Cruz, Mónica Aparicio González, Verónica Beatriz Rajal

## Abstract

The aim of this work was to evaluate if rivers could be used for SARS-CoV-2 surveillance to support health authorities. Five sampling points from three rivers (AR-1 and AR-2 in Arenales River, MR-1 and MR-2 in Mojotoro River, and CR in La Caldera River) from the Province of Salta (Argentina), two of them receiving the discharges of the wastewater plants (WWTP) of the city of Salta, were monitored from July to December 2020 during the first wave of COVID-19. Fifteen water samples from each point (75 samples in total) were collected and characterized physico-chemically and microbiologically and SARS-CoV-2 was quantitatively detected by RT-qPCR. In addition, two targets linked to human contributions, human polyomavirus (HPyV) and RNase P, were quantified and used to normalize the SARS-CoV-2 concentration, which was ultimately compared to the active reported COVID-19 cases. Statistical analyses allowed us to verify the correlation between SARS-CoV-2 and the concentration of fecal indicator bacteria (FIB), as well as to find similarities and differences between sampling points. La Caldera River showed the best water quality, and FIB were within acceptable limits for recreational activities. Although Mojotoro River receives the discharge of the northern WWTP of the city, it did not affect the water quality. Instead, the Arenales River presented the poorest water quality and the river at AR-2 was negatively affected by the discharges of the southern WWTP, which contributed to the significant increase of fecal contamination. SARS-CoV-2 was only found in about half of the samples and in low concentrations in La Caldera and Mojotoro Rivers, while it was high and persistent in the Arenales River. None of the two human tracers was detected in CR, only HPyV was found in MR-1, MR-2 and AR-1, and both were quantified in AR-2. The experimental and the normalized (using the two tracers) viral concentrations strongly correlated with the curve of active reported COVID-19 cases; thus, the Arenales River at AR-2 reflected the epidemiological situation of the city. This is, to the best of our knowledge, the first study that showed the dynamic of SARS-CoV-2 concentration in an urban river highly impacted by wastewater and proved that can be used for SARS-CoV-2 surveillance to support health authorities.

## 1. Introduction

The *Coronaviridae* family includes a broad spectrum of viruses that cause infections in various vertebrate animals including humans. Human Coronavirus are generally responsible for common colds. However, severe acute respiratory syndrome coronavirus 1 (SARS-CoV1) and Middle East respiratory syndrome coronavirus (MERS-CoV) were responsible for global epidemics, with high rates of morbidity and mortality, in 2003 and 2012, respectively (Peiris et al, 2003; Zaki et al, 2012). On 31 December 2019, the World Health Organization was informed about pneumonia cases of unknown origin in Wuhan, China. On 7 January 2020 the new virus, called 2019-nCoV at that moment, was identified as the cause and on 11 February 2020, it was named as the severe acute respiratory syndrome coronavirus 2 (SARS-CoV-2) by the International Committee on Virus Taxonomy (Gorbalenya et al., 2020). The virus spread rapidly among the population in the globe and the pandemic was declared in mid-March 2020. Since then, it has infected more than 176 million people and 3.8 million confirmed deaths by June 16^th^, 2021 (WHO, 2020).

SARS-CoV-2 is an enveloped single-stranded positive-sense RNA virus with an approximate diameter of 80 to 120 nm. The main transmission pathway is person-to-person, mainly airborne by aerosols, droplets, and sputum, formed during breathing, sneezing, coughing, or talking (Zhang et al., 2020a). Another way of transmission described initially was by contact with contaminated fomites; however, this hypothesis lost strength and is yet under discussion (Harrison et al., 2020; Kampf et al., 2020; Mukhra et al., 2020).

Although the 2019-nCoV infection (COVID-19) is mainly a respiratory disease, SARS-CoV-2 also replicates in the enterocytes from ileum and colon (Zhang et al., 2020b). Thus, a fraction of infected people, symptomatic or asymptomatic, excrete the virus in feces (Xiao et al., 2020b; Zhang et al., 2020c; Zhang et al., 2020d) and urine (Guan et al., 2020; Jeong et al., 2020) in variable concentrations. Furthermore, in one study the virus was isolated from urine (Sun et al., 2020) and in another from stool samples and propagated in Vero E6 cells (Xiao et al., 2020a). The fact that viral particles remain infective after being excreted through fecal matter, supports the hypothesis of a possible fecal-oral (Amirian, 2020; Jiang et al., 2020; Park et al., 2020; Wölfel et al., 2020) or a fecal-respiratory (Xiao et al., 2020a) route of transmission, although they have not been fully stablished yet.

The viral circulation in the population of a city, or a region, is reflected in the viral loads in their wastewater (Medema et al., 2020; Randazzo et al., 2020a). In fact, the so-called wastewater-based epidemiology has been applied in many developed countries (Ahmed et al., 2020; Lodder et al., 2020; Betancourt et al., 2021; Gonzalez et al., 2020; Albastaki et al., 2021; Saththasivam et al., 2021) to help authorities to better understand the situation and to make decisions regarding lockdowns. In most places, surveillance has been done in wastewater treatment plants, analyzing therefore, samples that represent the population of a city or part of it (Ahmed et al., 2020; Gonzalez et al., 2020; Kumar et al., 2020; Randazzo et al., 2020b; Rimoldi et al., 2020).

Rivers, as well as other water bodies, usually receive the discharges of treated wastewater (although sometimes with insufficient treatment) and, on some occasions, also (illegal) discharges of raw sewage. In this way, surface water can be impacted by wastewater. At the same time many of those aquatic environments are used as source water, for the irrigation of vegetables or for recreational purposes (Chávez-Díaz et al., 2019; Gutiérrez-Cacciabue et al., 2014; Poma et al., 2012). Previous studies about the microbiological quality of surface water in Salta, northwest of Argentina, demonstrated that fecal contamination was considerable in some rivers (Chavez-Díaz et al., 2020; Cruz et al., 2012; Gutierrez-Cacciabue et al., 2014), and multiple pathogens, including bacteria, enteric virus, and parasites, were found (Pisano et al., 2018; Poma et al., 2012; Prez et al., 2020), representing a risk for the population in contact with those waters (Poma et al., 2019).

The aim of this work was to evaluate if rivers could be used for SARS-CoV-2 surveillance to support health authorities. Three rivers from the province of Salta (northwest of Argentina), two of them receiving the discharges of the two wastewater plants of the city of Salta (main city in the province, 700,000 inhabitants), were monitored from July to December 2020 during the first wave of COVID-19. Water samples were physico-chemically and microbiologically characterized, and SARS-CoV-2 was quantitatively detected. In addition, two targets linked to human contributions were quantified and used to normalize the SARS-CoV-2 concentration, which was ultimately compared to the reported COVID-19 cases.

## 2. Materials and methods

### 2.1. Sampling sites

Three rivers from the province of Salta, northwest of Argentina, were selected for the study: Arenales, Mojotoro, and La Caldera Rivers (Figure 1). The first two are impacted by the discharges of the two wastewater treatments plants (WWTP) of the city of Salta (capital of the province, with 700,000 inhabitants) and vicinities, while the third one is used as source water upstream and as a recreational ambient downstream.

**Figure 1.**
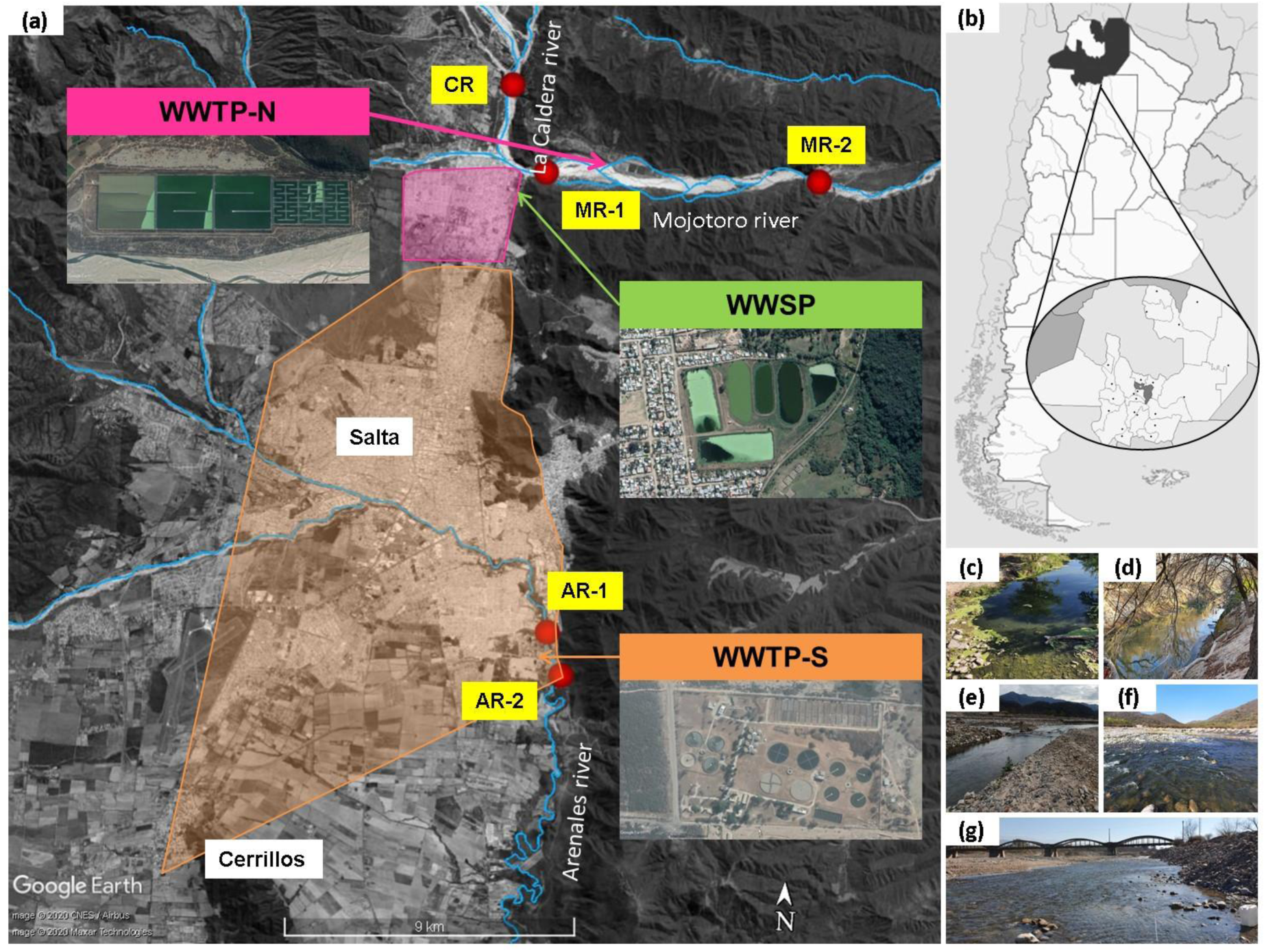
Geographical representation of the sampling sites (a) in Salta city and vicinities at the Province of Salta, northwest of Argentina (b). Three rivers were monitored: Arenales River before (AR-1) and after (AR-2) the discharge of the southern wastewater treatment plant (WWTP-S); Mojotoro River before (MR-1) and after (MR-2) the discharge of the northern wastewater treatment plant (WWTP-N), and La Caldera River (CR). An old wastewater stabilization pond (WWSP), out of use, located between CR and MR-1is also indicated. Shaded pink area corresponds to the coverage of WWTP-N, while shaded orange area corresponds to the coverage of WWTP-S. Illustrative photographs of the sampling points are shown: AR-1 (c), AR-2 (d), MR-1 (e), MR-2 (f), and CR (g).

The Arenales River (AR) is the main river that crosses the city of Salta, from west to southeast, that belongs to the Juramento-Salado watershed in the province of Salta, northwest of Argentina. While passing across the city, the river receives stormwater, industrial effluents, and illegal raw sewage (Poma et al., 2012; Lomniczi et al., 2007). Two sampling points were selected (Figure 1 a): AR-1 (Figure 1 c) and AR-2 (Figure 1 d) located 0.7 km upstream and 0.7 km downstream the southern wastewater treatment plant (WWTP-S) and the municipal landfill, respectively. Despite the chemical and microbiological contamination of this river (Lomniczi et al., 2007; Poma et al., 2012; Prez et al., 2020), it is used for domestic irrigation of vegetables and for recreational purposes.

La Caldera River (CR) is located 13 km north from the city of Salta. It is used as source water and for recreational activities. Indeed, it is the most visited river in Salta citýs vicinities because of its landscapes and accessibility. One sampling site (CR) in a recreational area was selected for this study (Figure 1 e).

The Mojotoro River (MR) is originated from the convergence of La Caldera and Vaqueros Rivers (Figure 1 a). It represents a natural border between Salta city and vicinities, going from urban to semi-rural areas, and it is mainly used for recreation and gravel extraction. Two sampling points, 10 km away from each other, were selected for monitoring: MR-1 (Figures 1 a, e) and MR-2 (Figures 1 a, f), located 5 km before and 5 km after the discharge of the northern wastewater treatment plant (WWTP-N) (Figure 1 a), respectively. There is also an old, out of use, wastewater stabilization pond (WWSP) located 3.5 km after CR and 0.7 km before MR-1 (Figure 1 a).

### 2.2. Sampling and characterization of water samples

A total of 15 monitoring campaigns were carried out weekly initially and then every other week, from the 13 July to 28 December 2020. Water samples were characterized physico-chemical and microbiologically. Physico-chemical variables: pH, temperature (°C), turbidity (NTU), and conductivity (mS/cm), were measured in situ using a multiparametric probe U10 Horiba (Japan). For microbiological characterization, one-liter water samples were collected in sterile bottles and refrigerated until fecal indicator bacteria (FIB) were analyzed in the laboratory. Also 20-liter water samples were collected in clean and rinsed plastic carboys at each point and brought to the laboratory for concentration and molecular analysis.

Total (TC) and thermotolerant (TTC) coliforms were determined by the multiple-tube fermentation method, culturing them on MacConkey broth (Britania, Argentina) at 37 and 44.5 °C for 48 h, respectively (Eaton et al., 2005). *Escherichia coli* (EC) and enterococci (EN) were enumerated through the Membrane Filter technique (Eaton et al., 2005). *E. coli* was cultured in modified mTEC Agar (Fluka, USA) at 35 °C for 2 h and 44.5 °C for 22 h (Method 1603, USEPA 2002a), and enterococci in mE Agar (Difco, USA) at 41 °C for 48 h and confirmation in Esculin-Iron Agar at 41 °C for 20 min (Method 1106.1, USEPA 2002b). Results for TC and TTC were expressed as Most Probable Number (MPN) per 100 mL, while for EC and EN they were expressed as Colony Forming Unit (CFU) per 100 mL.

### 2.3. Water concentration and nucleic acid extraction

Twenty-liter water samples were concentrated down to 80 mL by ultrafiltration using a disposable hemodialysis hollow fiber cartridge (FX Classix 100, Fresenius Medical Care, Germany), following a previously optimized and validated protocol for the detection of enteric viruses (Poma et al, 2012; Rajal et al, 2007). A fraction of the final concentrated water sample (140 µL) was employed for nucleic acid extraction using a commercial kit (Puro Virus RNA, Productos Bio-Lógicos, Argentina), producing 50 µL of extract which was used for the quantitative detection of SARS-CoV-2, Polyomavirus (HPyV) and Ribonuclease P (RNase P).

### 2.4. Quantitative detection of SARS-CoV-2

The detection of SARS-CoV-2 was performed on the RNA extracts from each concentrated water sample by RT-qPCR based on the detection of a region of the N gene (N1 system) validated by the US Centers for Disease Control and Prevention (CDC, 2020).

The determination and quantification of the gene N was performed in a StepOne Plus real time PCR system (Applied Biosystem). The final 20 µL reaction volume contained 5 µL of TaqMan Fast virus 1-Step Master Mix 4X (Applied Biosystems), primers and probe solutions, water (to complete 15 µL), and 5 µL of template. The final concentrations of forward and reverse primers were 400 nM each while it was 250 nM for the probe. The RT-qPCR was initiated with reverse transcription at 50 °C for 5 minutes followed by a step of 95 °C for 20 seconds and 45 cycles of 95 °C for 15 seconds and 59 °C for 1 minute.

Dilutions of the RNA extract were performed when needed to overcome inhibition. Reactions were performed in duplicate, simultaneously with a non-template control (MiliQ water) and DNA plasmid (nCoV-ALL-Control Plasmid, Eurofins) as positive control. A protocol for rapid digestion with EcoR1 (Promega, USA) was used to linearize the plasmid, following the manufacturer’s instructions.

The standard curve for SARS-CoV-2 quantification (y = -3.4700 x + 43.1616; R^2^= 0.9820; amplification efficiency: 94%; dynamic range: 8) was performed with eight 10-fold serial dilutions, in triplicate, of the DNA plasmid that contains the interest target.

### 2.5. Normalization with Human Polyomaviruses (HPyV) and Ribonuclease P (RNase P) as human input indicators

The viral concentration was expected to show variations along time. These variations could be due to changing viral discharges into the rivers but also due to fluctuations in the flow rate, i.e. because of stormwater. Thus, to accurately assess the evolution of SARS-CoV-2 concentration, without the impact of external factors, two human indicators (HPyV and RNaseP) were used as normalizers.

Human JC (JCPyV) and BK (BKPyV) polyomaviruses were quantitively detected by qPCR using the oligonucleotides and reaction conditions described by Barrios et al. (2018). On the other hand, a fragment of the gene encoding for the human Ribonuclease P (RNase P) was quantitively detected by qPCR using the system described by the US Centers for Disease Control and Prevention (CDC, 2020).

The final volume of each qPCR reaction contained 15 µL of reaction mix containing SensiFAST Probe HiRox kit 2X (Bioline) and specific primers and probes to reach the final concentrations established (a summary of all the oligonucleotides used for all the targets, including those for the detection of SARS-CoV-2 is shown in Table 1) and 5 µL of template. Cycling parameters were 95 °C for 5 minutes for polymerase activation and 45 cycles of 95 °C for 10 seconds for denaturation and 60 °C for 50 seconds for annealing/extension.

**Table 1.**
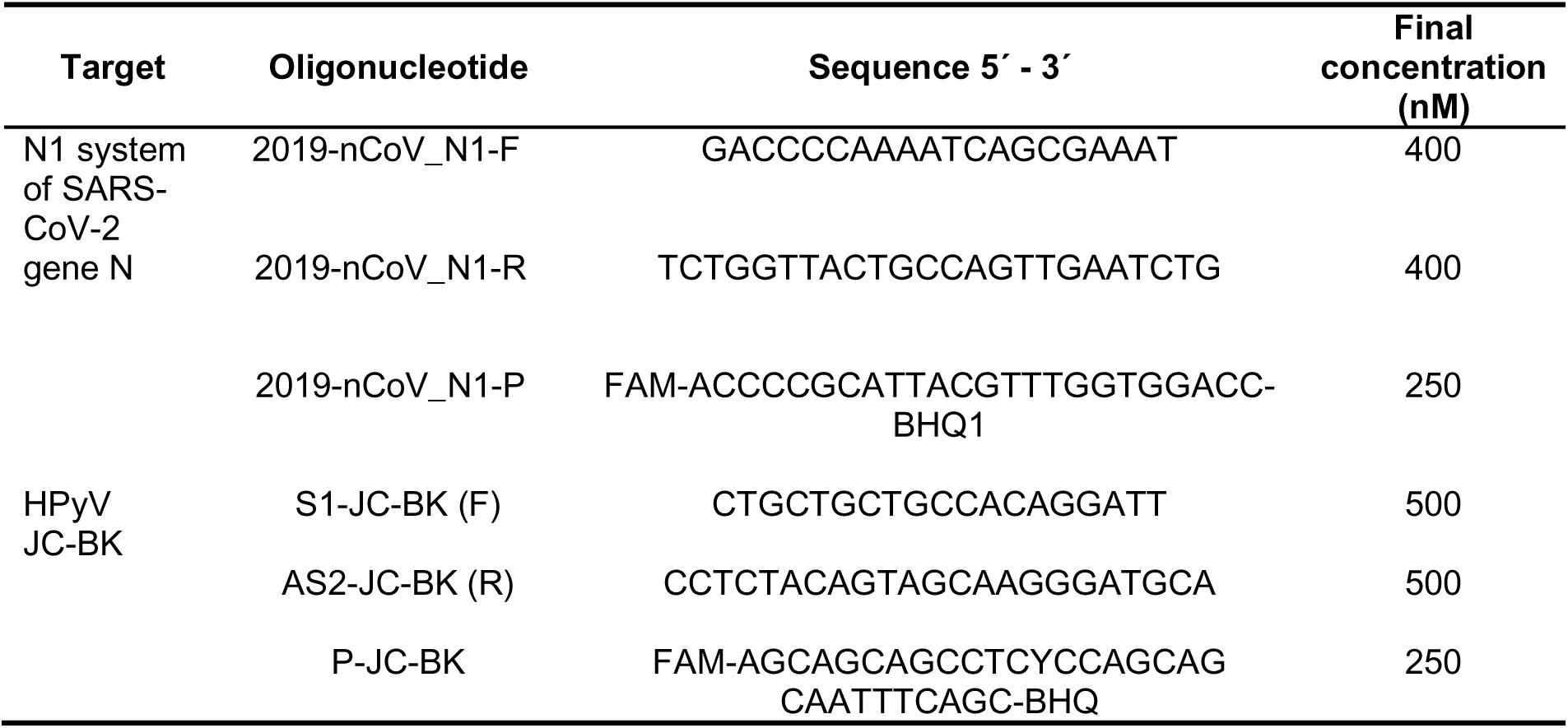
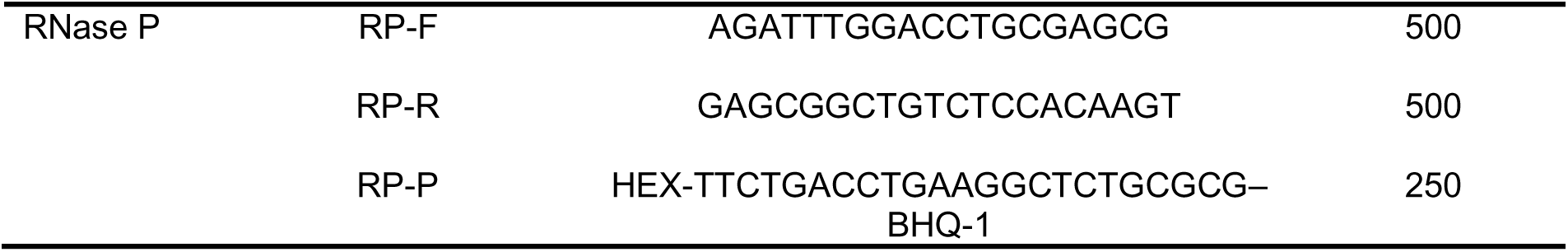
Oligonucleotides: forward (F) and reverse (R) primers and Taq-Man® probes (P) used for the detection of SARS-CoV-2 N gene (CDC, 2020), HPyV (Barrios et al., 2018) and RNase P gene (CDC, 2020).

Standard curves for HPyV (y = -3.656 x + 39.317; R^2^= 0.996; amplification efficiency: 88%; dynamic range 8) and RNase P (-3.71 x + 42.8280; R^2^= 0.9991; dynamic range 7) markers, were built using 10-fold dilutions of the corresponding plasmid DNA, containing the target sequences, in triplicate.

All the extracted nucleic acids from the concentrated water samples were analyzed for HPyV and RNaseP, by duplicate, using positive (plasmid) and negative (ultrapure water) controls. These results were used for the normalization of the SARS-CoV-2 concentrations; three alternative normalizations were assessed, as follows:

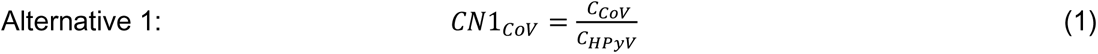

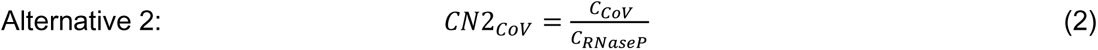

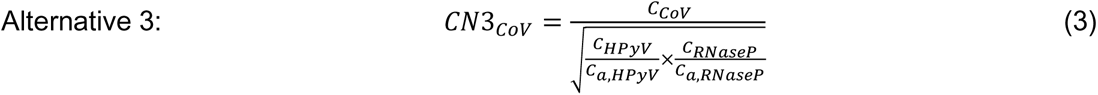

Where *C* (gc/mL) is the concentration and subindexes *CoV* (as for SARS-CoV-2), *HPyV* and *RNaseP* indicate the targets; *C_a_* (gc/mL) is the average concentration and *C_N_* (gc/mL) is the normalized concentration. The super index (*i*) (*i* = 1, 2, 3) indicates the normalization alternative.

### 2.6. COVID-19 cases

Reported COVID-19 cases for the city of Salta were compiled and the prior two-weeks accumulated amount were assigned to each monitoring date to compare the epidemiological curve to the evolution of the viral concentration in the Arenales River. The reason for choosing this river was that it crosses the city receiving the impact of various human activities (domestic and industrial) and also, it receives the discharges of the main wastewater treatment plant (WWTP-S) (covers 85% of the population), which is outdated and not sufficient for treating the volume of a fast-growing city.

### 2.7. Statistical analyses

Physico-chemical variables (temperature, pH, conductivity, and turbidity) measured along the monitoring campaigns were compiled to show descriptive statistics: mean and standard deviation, median and range of variability (minimum – maximum).

To assess data variability during the monitoring campaign within each river and regarding the impact due to SARS-CoV-2, different statistical tests were run.

Most analyses were carried out using non-parametric tests, due to the non-normal distribution of the data (*p* < 0.001). Only the correlation between concentrations of HPyV and RNase P was carried out with a parametric test (Pearson) as both datasets showed normal distribution. Normality was tested by the Shapiro–Wilks W-test (Shapiro and Francia, 1972).

Kruskal-Wallis test (non-parametric) was performed with the physico-chemical and microbiological variables measured to compare all the sampling points. Different letters were assigned when there were significant differences within the variables between the sampling points with *p*-values < 0.05.

Correlations between all the physico-chemical variables, and fecal indicator bacteria and SARS-CoV-2 concentrations measured for all the sampling points were assessed by Spearman test (non-parametric). This test was also applied to all the same variables for each sampling point. Correlations were considered significant when the probabilities (*p*) were lower than 0.05 and they were arbitrary considered weak when coefficients were lower than 0.40, moderate when were between 0.40 and 0.75, and strong when were higher than 0.75.

A Cluster Analysis (CA) and a Linear Discriminant Analysis (DA) were applied using nine variables (TC, TTC, EC, EN, SARS-CoV-2, T, conductivity, pH and turbidity) to assess the behavior of the data in a multivariate sense. The CA aimed to search similarities between sampling points. This multivariate statistical test was performed by the Ward Method. Euclidean distances were calculated for each case. The DA was applied to distinguish between sampling points. The goal is to discriminate two or more groups of objects by maximizing their differences (Di Renzo et al., 2016). All the variables were standardized before the multivariate performance, to eliminate the effect of scale.

Statistical analyses were run with the InfoStat software (Di Rienzo et al., 2016) and RStudio version 4.0.0 (Ihaka and Gentleman, 1996; R Development Core Team, 2005; RStudio Team, 2020; Wickham, 2009).

Finally, the parametric Pearsońs test was performed to evaluate is there was correlation between the concentrations of HPyV and RNaseP. In addition, Spearman test was performed to assess for correlation between the evolution of the concentration of SARS-CoV-2 in the Arenales River, at AR-1 and AR-2, and the number of active reported cases in Salta city. Then the test was applied for the normalized SARS-CoV-2 concentration at AR-2 and the number of cases.

## 3. Results

A total of 15 sampling events were carried out from July to December of 2020 in three rivers, monitoring a total of five points: AR-1 and AR-2 at the Arenales River, CR at La Caldera River, and MR-1 and MR-2 at Mojotoro River.

### 3.1. Physico-chemical characterization of water samples

Four physico-chemical variables (temperature, pH, conductivity, and turbidity) were measured in water (Table 2). Water temperature (7.2 - 23.8 °C) was directly affected by environmental temperature (5.0 - 25.0 °C) (July to middle September is winter and middle September to middle December is spring in the South Hemisphere), as expected.

**Table 2.** Physico-chemical variables measured in surface water, along 15 sampling events, in the Arenales River at AR-1 and AR-2 (before and after the discharges of the southern wastewater treatment plant, WWTP-S, respectively; distance between AR-1 and AR-2 is 1.4 km and WWTP-S is separated 0.7 km from each), in La Caldera River (CR), and in the Mojotoro River at MR-1 (after an old wastewater stabilization pond and before the discharges of the northern wastewater treatment plant, WWTP-N) and MR-2 (after the discharges of the WWTP-N; distance between MR-1 and MR-2 is 10 km and WWTP-N is 5 km away from each). K-W: Bonferroni Post Hoc test from Kruskal Wallis. Medians with different letters are significantly different (*p <* 0.05).

| Variables | Sampling points | Mean ± SD | Median | Range (min-max) | K-W |
| --- | --- | --- | --- | --- | --- |
| Temperature (°C) | AR-1 | 17.41 ± 3.99 | 16.30 | 12.10 - 23.50 | AB |
| | AR-2 | 19.23 ± 3.19 | 18.40 | 14.70 - 23.80 | A |
| | CR | 15.83 ± 4.73 | 14.30 | 7.20 -23.80 | C |
| | MR-1 | 15.69 ± 4.18 | 15.00 | 8.00 - 22.50 | A |
| | MR-2 | 16.28 ± 4.88 | 16.00 | 7.80 - 23.80 | BC |
| pH | AR-1 | 8.07 ± 0.27 | 8.12 | 7.64 - 8.54 | E |
|  | AR-2 | 7.90 ± 0.12 | 7.91 | 7.70 - 8.09 | E |
|  | CR | 8.45 ± 0.28 | 8.46 | 7.89 - 8.82 | D |
|  | MR-1 | 7.96 ± 0.40 | 7.94 | 6.99 - 8.90 | D |
|  | MR-2 | 8.24 ± 0.32 | 8.24 | 7.32 - 8.63 | D |
| Conductivity<br>(mS/cm) | AR-1 | 0.46 ± 0.07 | 0.48 | 0.31 - 0.54 | G |
|  | AR-2 | 0.58 ± 0.07 | 0.59 | 0.41 - 0.74 | G |
|  | CR | 0.21 ± 0.03 | 0.20 | 0.17 - 0.27 | F |
|  | MR-1 | 0.24 ± 0.01 | 0.24 | 0.21 - 0.27 | F |
|  | MR-2 | 0.25 ± 0.02 | 0.24 | 0.20 - 0.28 | F |
| Turbidity<br>(NTU) | AR-1 | 17.07 ± 32.51 | 8.00 | 4.00 - 134.00 | H |
|  | AR-2 | 88.27 ± 28.44 | 80.00 | 55.00 - 159.00 | J |
|  | CR | 16.07 ± 13.78 | 13.00 | 5.00 - 56.00 | HI |
|  | MR-1 | 11.47 ± 5.57 | 10.00 | 5.00 - 26.00 | HI |
|  | MR-2 | 16.27 ± 5.36 | 16.00 | 5.00 - 26.00 | I |

Although the maximum temperatures were similar for all the rivers, the minimum values were higher for AR-1 and AR-2 (Arenales River is in the middle of the city) compared to those from CR, MR-1 and MR-2 which are in the vicinities of Salta, in open space and more exposed to the wind and elements.

Kruskal-Wallis test was performed with physico-chemical variables measured in the five sampling points analyzed. The sampling points CR, MR-1 and MR-2 were similar except for the temperature that was different in MR-1. The two monitoring points at the Arenales River (AR-1 and AR-2; distance was 1.4 km, the WWTP-S was 0.7 km away from each) only showed significant differences in the turbidity, which was much higher in AR-2, probably due to the discharges of the WWTP-S, also reported in a previous work (Poma et al, 2012).

### 3.2. Bacterial characterization

The highest concentrations of all FIBs were found in the Arenales River, followed by Mojotoro River and La Caldera River (Figure 2). In general, variability in bacteria concentrations (within two orders of magnitude) was lower in AR-1 and AR-2 than in the other sampling points (Figure 2). Conversely, the concentrations of total and thermotolerant coliforms were highly variable for MR-1, MR-2, and CR.

**Figure 2.**
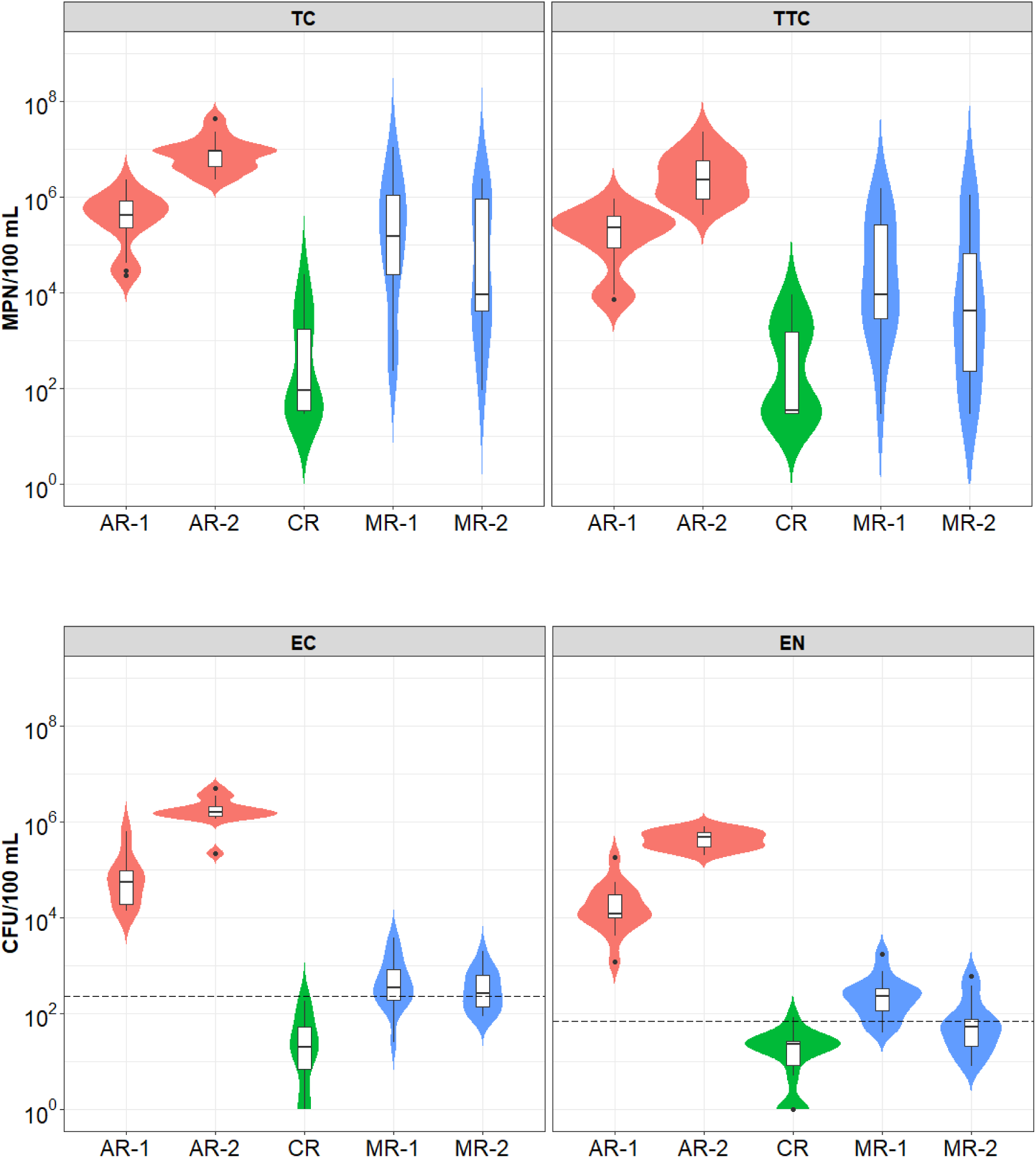
Violin plots representing indicator bacteria determined in 15 monitoring events in five different sampling points (75 samples in total): AR-1 and AR-2 in the Arenales River (before and after the discharges of the southern wastewater treatment plant, respectively), CR in La Caldera River, and MR-1 and MR-2 in the Mojotoro River (before and after the discharges of the northern wastewater treatment plant, respectively). The concentrations of total (TC) and thermotolerant coliforms (TTC) are expressed as the Most Probable

Bacteria concentrations in Arenales River exceeded 10^4^ CFU or MPN per 100 mL and furthermore, they surpassed the acceptable limits for *E. coli* and enterococci (235 CFU/100 mL and 71 CFU/100 mL, respectively, USEPA 2012) in all the samples analyzed. The point AR-2, 700 m downstream from AR-1, was highly impacted by fecal contamination of untreated or insufficiently treated wastewater effluents from the WWTP-S (*p* < 0.05 for all the FBI) (Table 3).

**Table 3.** Bonferroni Post Hoc test from Kruskal Wallis for fecal indicator bacteria (total and thermotolerant coliforms, *E. coli*, and enterococci) determined in five sampling points: CR in La Caldera River, MR-1 and MR-2 in Mojotoro River, and AR-1 and AR-2 in Arenales River. Medians with different letters for each variable are significantly different (*p <* 0.05).

|  | Total<br>coliforms | Thermotolerant<br>coliforms | *E. coli* | Enterococci |
| --- | --- | --- | --- | --- |
| CR | A | A | A | A |
| MR-1 | A | A | B | B |
| MR-2 | A | A | B | AB |
| AR-1 | B | B | C | C |
| AR-2 | B | C | C | C |

Regarding Mojotoro River, bacteria concentrations were similar in both sampling sites (Table 3) showing that the effluents of the WWTP-N have no impact on the microbial quality of water. However, the concentration of *E. coli* and enterococci exceeded the recreational limits in most of the samples (67 and 80% for MR-1, respectively, and 53 and 33% for MR-2, respectively). In contrast, La Caldera River showed the best water quality, appropriate for recreational activities. In fact, all the samples analyzed (15 in total) were within the accepted values for *E. coli*; however, one of them exceeded the limit for enterococci.

Number per 100 mL, while those of *E. coli* (EC) and *Enterococcus* sp. (EN) are in Colony Forming Units (CFU) per 100 mL. Violin shapes show the data kernel probability with the boxplots embedded. Boxplots show the interquartile range (IQR) in boxes divided by median values (horizontal lines) with whiskers depicting ±1.5 IQR and outliers as points. The segmented horizontal lines represent the limit values accepted by legislation for recreational water (USEPA, 2012).

### 3.3. Quantification of SARS-CoV-2, HPyV and RNase P

All the samples (75 in total) collected during the 15 sampling events (from July to December 2020, during the first wave of COVID-19) at five different points in three rivers, were concentrated, the nucleic acids were extracted, and then analyzed for SARS-CoV-2, HPyV and RNase P (Figure 3). In La Caldera River, from the three targets evaluated only SARS-CoV-2 was found in around half of the samples (53.3% samples were non-detects) and in those cases the viral concentrations were lower than 10^5^ gc/L. The same occurred in the Mojotoro River where 7 and 6 out of 15 (46.6 and 40.0%, respectively) samples were non detects at MR-1 and MR-2, respectively. However, HPyV was also detected in that river in both sampling points, in 14 and in 10 (out of 15) samples at MR-1 and MR-2, respectively. The HPyV was detected in all the samples in Arenales River, in both sampling points, showing the impact of human contamination. This effect was also seen through the detection of RNase P that was only found in AR-2 samples. While SARS-CoV-2 was not detected during the first five monitoring campaigns in AR-1, it was quantified in all samples collected at AR-2. The two targets selected for tracing human contamination showed a similar behavior (coefficient was 0.7228; *p* = 0.0021; positive strong correlation) along time. However, the concentrations of HPyV were around one order of magnitude higher than those of RNase P, which may be the reason for RNase P not being found consistently in other sites (average ratio HPyV/RNaseP was 15.8 ± 6.4, without considering one outlier on 27 July, where the ratio was 299.3).

**Figure 3.**
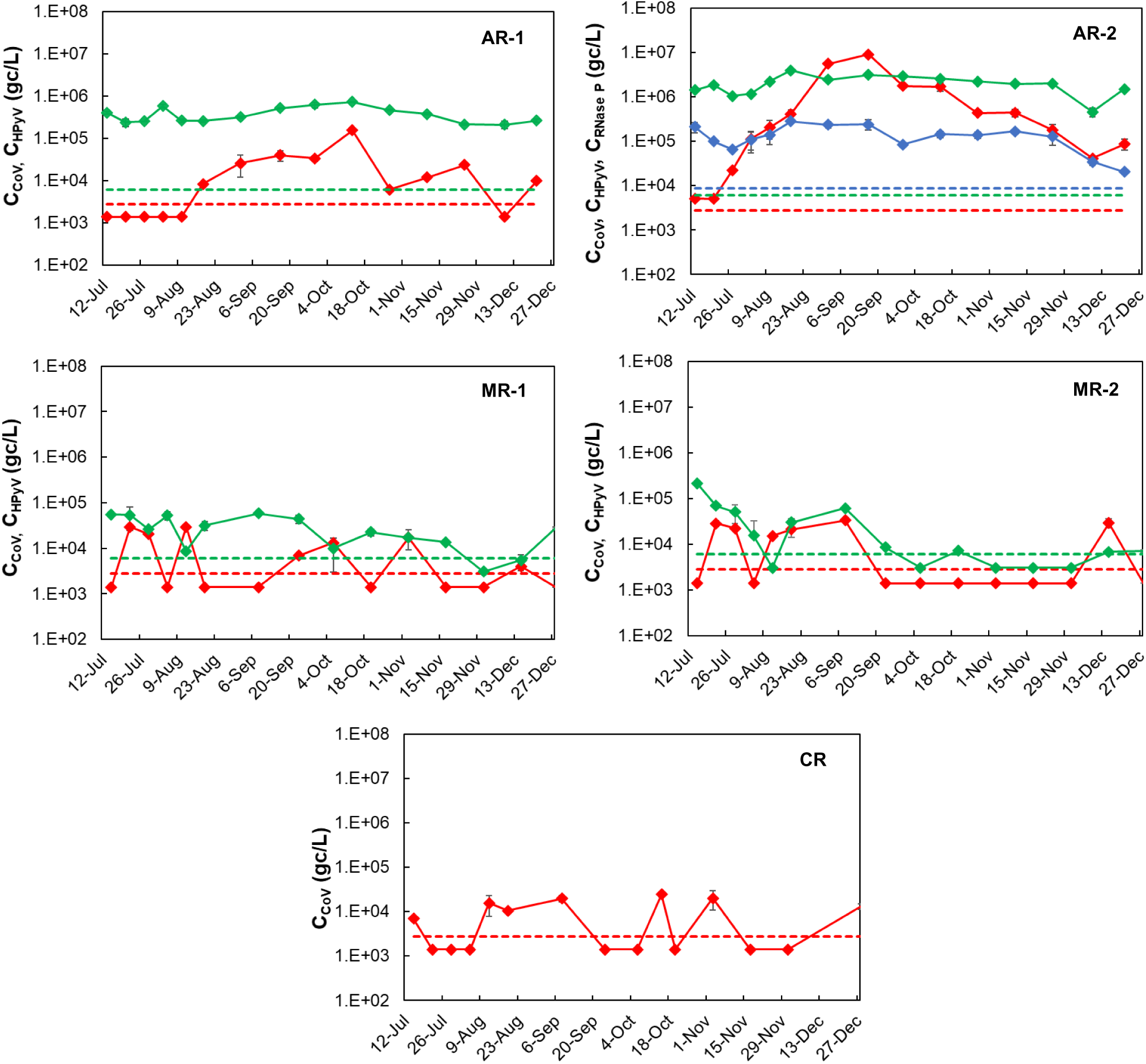
Evolution of SARS-CoV-2 (C_CoV_, red solid line), human polyomavirus (C_HPyV_, green solid line), and RNase P (C_RNase P_, blue solid line) concentrations, determined in five sampling points: AR-1 and AR-2 in Arenales River, MR-1 and MR-2 in Mojotoro River, and CR in La Caldera River, from July to December 2020 (15 sampling events). Dashed lines represent the limit of detection (LOD) for SARS-CoV-2 (red), HPyV (green) and RNase P (blue). Non detects were arbitrarily represented as LOD/2. Values plotted are the average of duplicates and error bars correspond to the standard deviation (some bars are too small to be seen). Concentrations are expressed in gene copies per liter (gc/L).

### 3.4. Relationship between water quality and SARS-CoV-2 concentration

Spearman correlation was evaluated using all the physico-chemical, FIBs and SARS-CoV-2 concentration data, including all the sampling points (Supplementary Information, Table S1). Temperature was the only variable that did not show any correlation. Positive strong correlations were found between pH and COND, TURB and FIBs, and among FIBs. Regarding CoV, also positive strong correlations were confirmed with pH and COND, while they were moderate with TURB and all FIBs (Supplementary Information, Table 1).

When Spearman correlation was performed for each sampling point instead, positive strong or moderate correlations were found (*p* < 0.05) between COND and pH (and with temperature for MR-2 only) and also between TC, TTC and EC for all the sampling points (Table 4). In all cases, except for MR-2, there were positive moderate correlations (*p* < 0.05) between TURB and at least two FIBs. Only for AR-1 and AR-2 positive moderate correlations (*p* < 0.05) were found between CoV and pH, CoV and TURB, and EN and another FIB (Table 4).

**Table 4.**
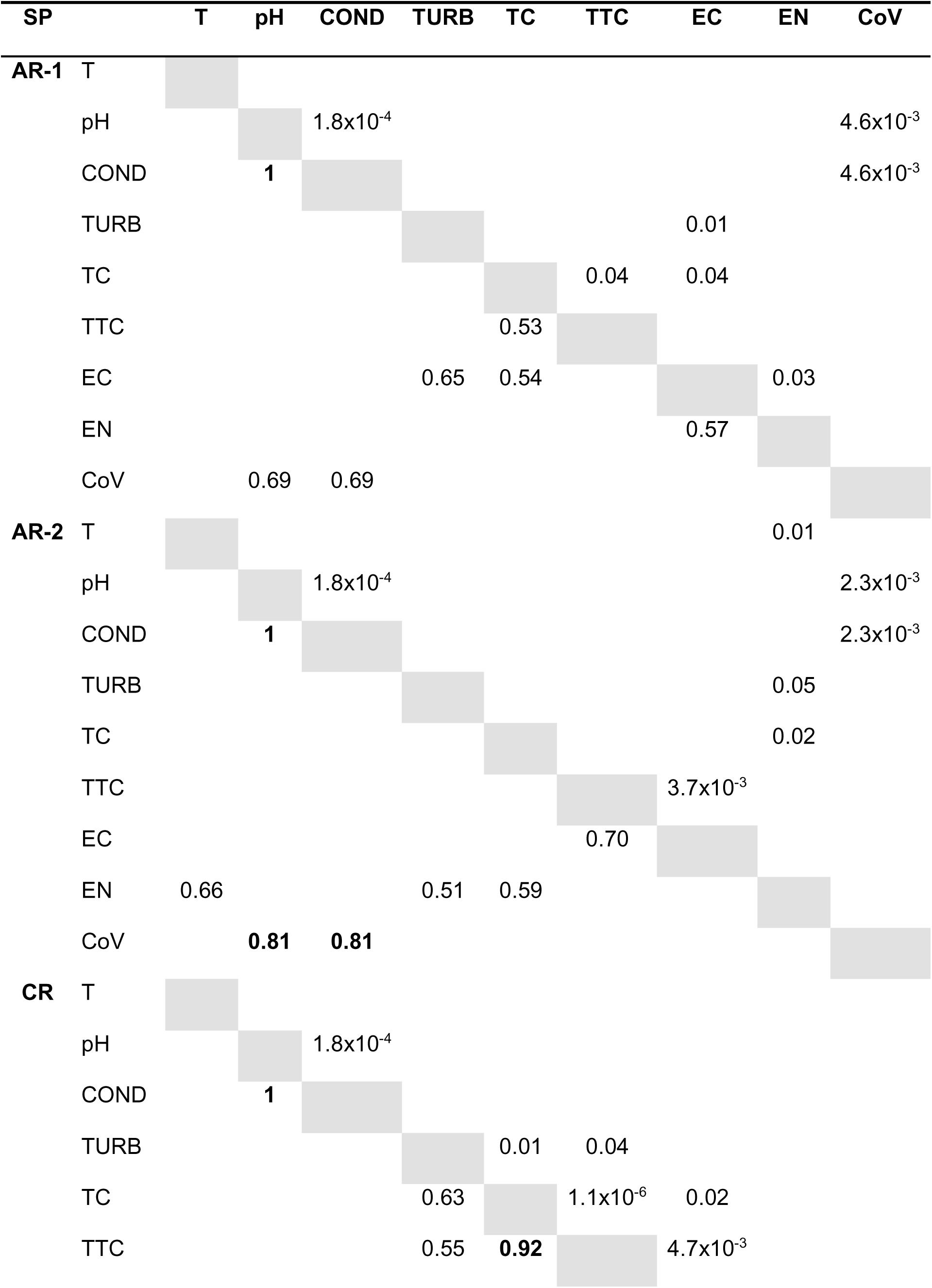
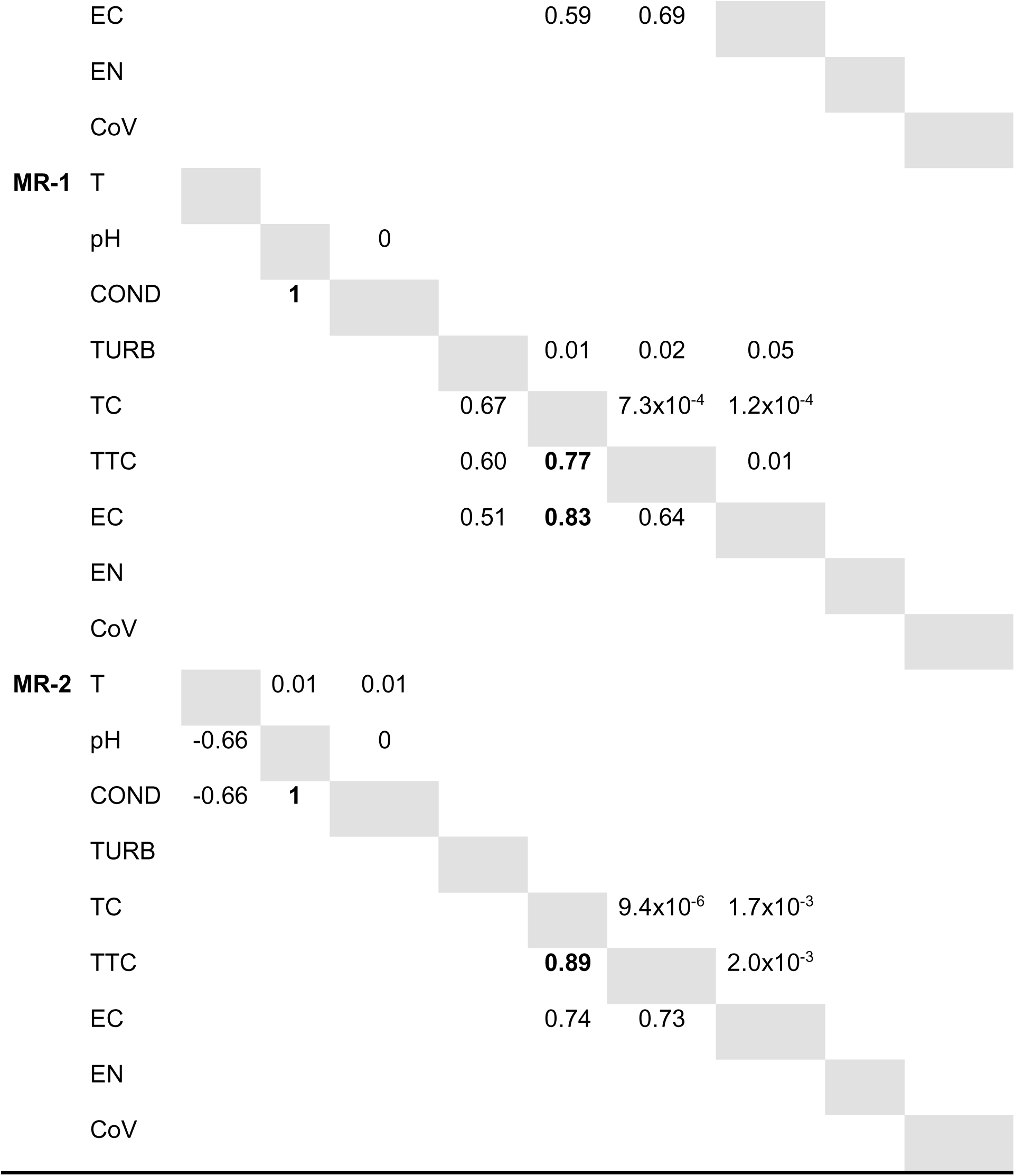
Coefficients and probabilities (below and above the main diagonal, respectively) from Spearman correlation test using the physico-chemical variables, fecal indicator bacteria and SARS-CoV-2 concentration (CoV) for each sampling point (SP): AR-1 and AR-2 in Arenales River; CR in La Caldera River; MR-1 and MR-2 in Mojotoro River. Only significant correlations (*p* < 0.05) are shown and those that are strong are indicated in bold. Physico-chemical variables: temperature (T), pH, conductivity (COND), turbidity (TURB). Fecal indicator bacteria: total coliforms (TC), thermotolerant coliforms (TTC), *E. coli* (EC), enterococci (EN).

Cluster analysis including all the variables measured for a total of 75 water samples from five sampling points was performed and the Euclidean distances were determined (cophenetic correlation was 0.906) (Figure 4). The most similar sampling points were MR-1 and MR-2 (Euclidean distance was 0.69), verifying once again that the Mojotoro River was not negatively impacted by the discharges of the WWTP-N. Conversely, the most different sampling points were CR and AR-2 (Euclidean distance was 6.99), in agreement with other observations, that CR was the point with the best water quality and AR-2 the most impacted one by the discharges of the WWTP-S.

**Figure 4.**
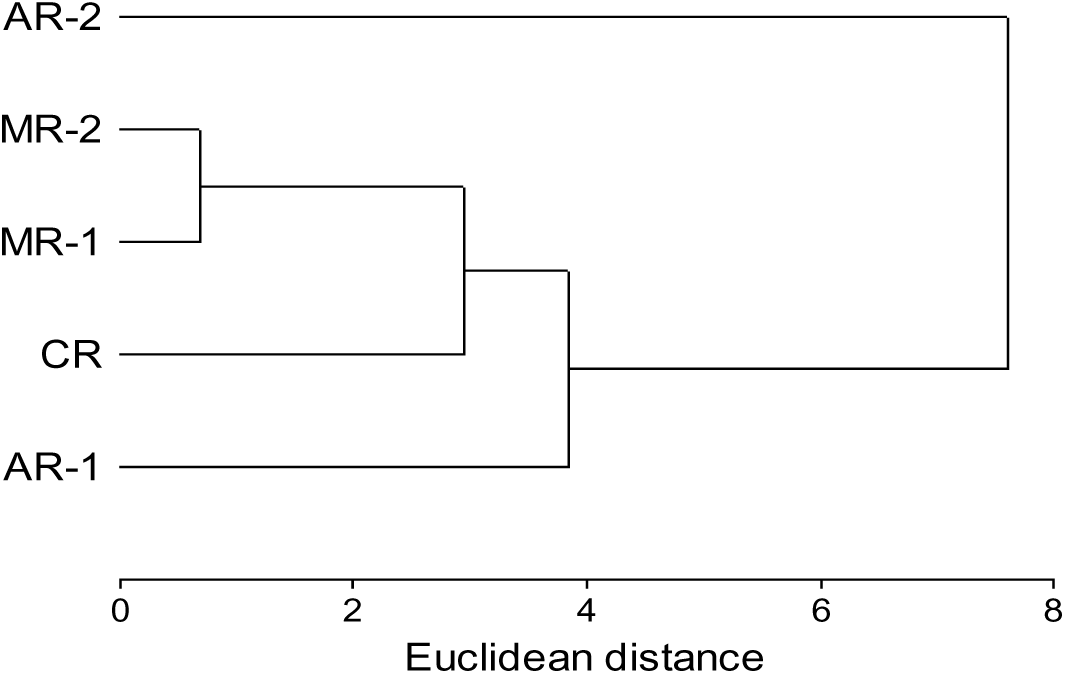
Cluster analysis performed with nine variables (temperature, pH, conductivity, turbidity, and concentrations of total and thermotolerant coliforms, *E. coli*, enterococci and SARS-CoV-2) measured in a total of 75 water samples collected in five sampling points: AR-1 and AR-2 in the Arenales River, MR-1 and MR-2 in the Mojotoro River, and CR in La Caldera River, from July to December 2020 (15 sampling events).

The discriminant analysis performed with nine the variables measured in 75 water samples along 15 sampling events showed two main groups (Figure 5). At the right side there were those samples from the Arenales River, where AR-1 was also discriminated from AR-2, due to the high microbial contamination. The second group, at the left side, included the samples from Mojotoro River, with MR-1 and MR-2 mingled together, slightly separated from those from Caldera River, which was the one with best microbial water quality.

**Figure 5.**
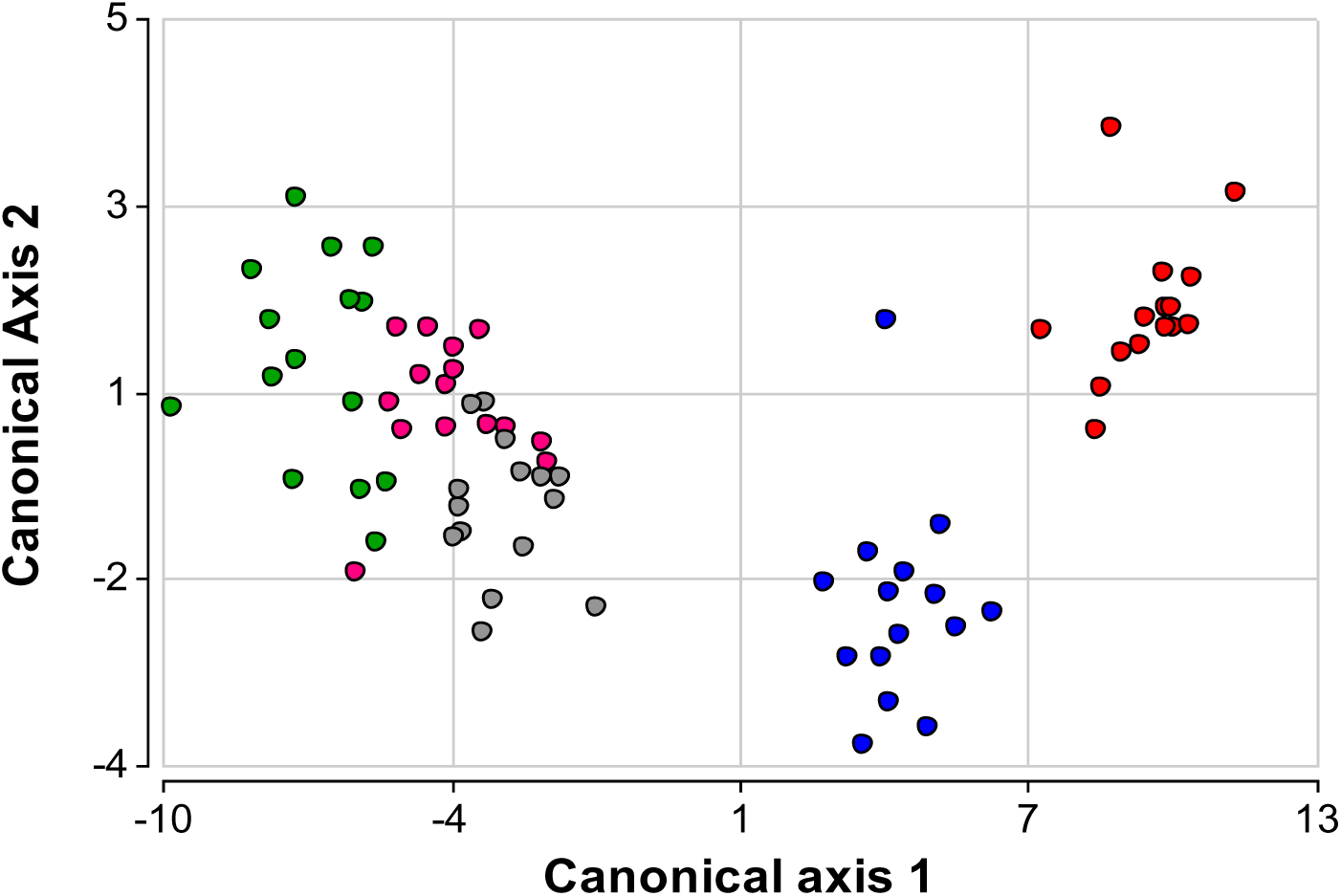
Plots of the first two axes (with eigenvalues higher than 1, explaining 97.7% of the accumulated variance) obtained with a linear discriminant analysis conducted with nine variables: temperature, pH, conductivity, turbidity, and concentrations of total coliforms, thermotolerant coliforms, *E. coli*, enterococci, and SARS-CoV-2. A total of 75 water samples were collected in five sampling points: AR-1 (red circles) and AR-2 (blue circles) in the Arenales River, MR-1 (gray circles) and MR-2 (pink circles) in the Mojotoro River, and CR (green circles) in La Caldera River, from July to December 2020 (15 sampling events).

### 3.5. Comparison between SARS-CoV-2 concentration and reported COVID-19 cases

The total of COVID-19 reported cases in the city of Salta, accumulated for 14 days before each monitoring campaign, were assigned to that sampling date. The evolution of the number of cases in Salta city was confronted with the SARS-CoV-2 concentration found in the Arenales River (Figure 6). This was not performed for the Mojotoro and La Caldera Rivers as SARS-CoV-2 was detected in around half of the samples, following no pattern, in low concentration, while the number of cases were constantly increasing.

**Figure 6.**
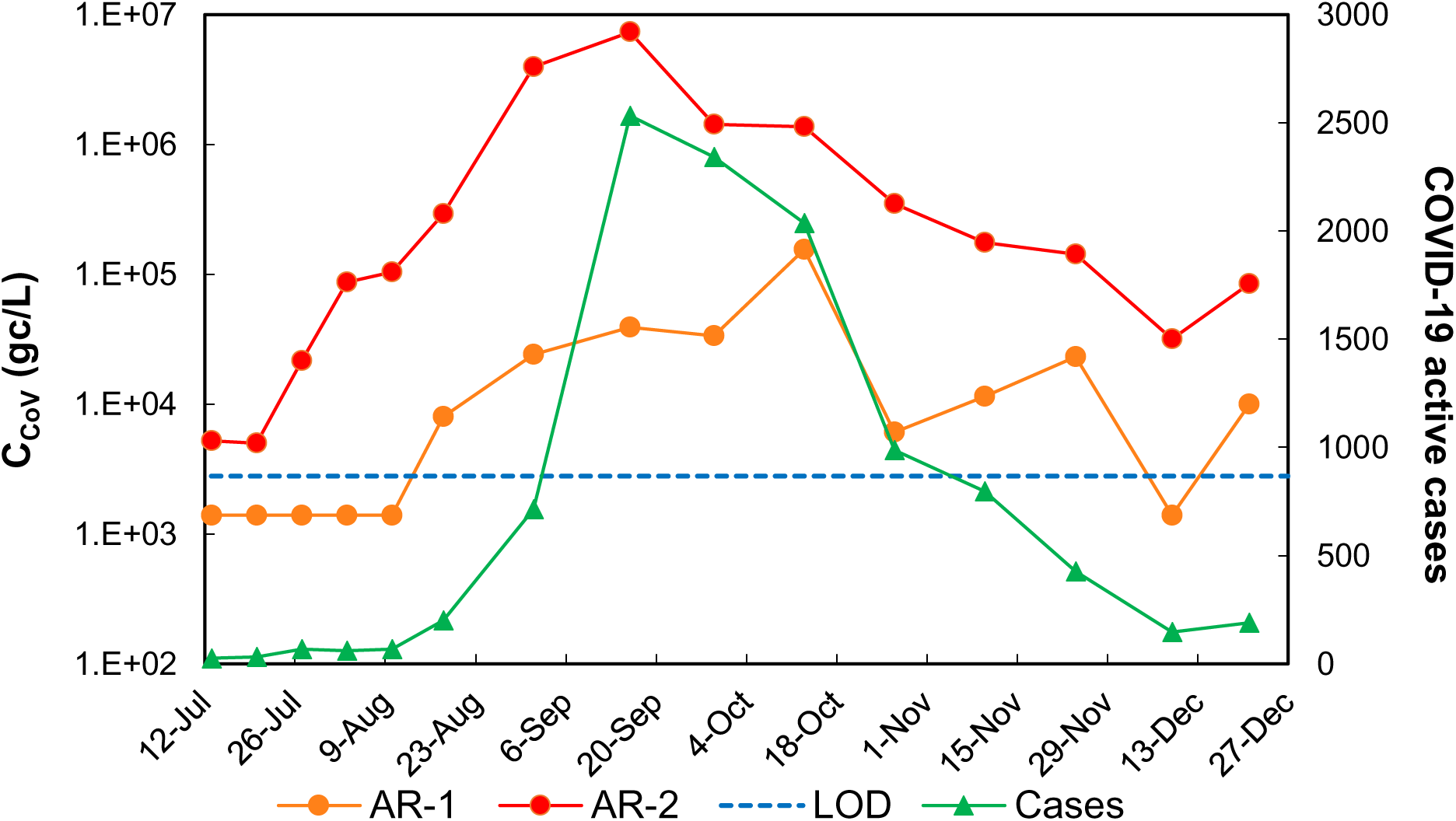
Evolution of SARS-CoV-2 concentration (C_CoV_) in the Arenales River at two sampling points: AR-1 and AR-2 before and after the southern wastewater treatment plant, respectively, and of the number of COVID-19 active cases accumulated for 14 days before sampling. LOD: limit of detection for SARS-CoV-2 in surface water. Non-detects were arbitrarily represented as LOD/2.

The viral concentration curve anticipated the epidemiological one. The ratio between the viral concentration and the number of active cases (14 days before) was variable, being 32 gc/mL/case the average for AR-1 and 1099 gc/mL/case the average for AR-2. The latest was a consequence of a higher fecal content in AR-2 due to the WWTP discharges in the river and was highly influenced by the extreme values. In fact, at the beginning and towards the end of the epidemiological curve the ratio was around 281 gc/mL/case while it was 2034 gc/mL/case during the two months (August-September) when the viral dissemination was higher.

Spearman correlation coefficient indicated a positive association between the number of cases and the viral concentrations in the Arenales River, at AR-1 (ρ = 0.89; *p* = 0.00001) and at AR-2 (ρ = 0.89; *p* = 0.00084).

### 3.6. Normalization of the SARS-CoV-2 concentration

The experimental concentration of SARS-CoV-2, although is calculated based on the detection of the gene of interest in water samples, is subject to fluctuations due to variations in the flow rate, i.e. from precipitations or from discharges of the wastewater plant. One strategy to compensate for that, thus, to help the data to get closer to the real situation, is to normalize the data, with some variable that accounts for the human input in this case. Two targets, T antigen for HPyV and RNase P, linked to human contamination were selected for that. Therefore, the concentration of SARS-CoV-2 in the Arenales River was normalized following three different alternatives, including the concentration of HPyV (Alternative 1), the concentration of RNase P (Alternative 2) and a combination of both (Alternative 3) (Figure 7).

**Figure 7.**
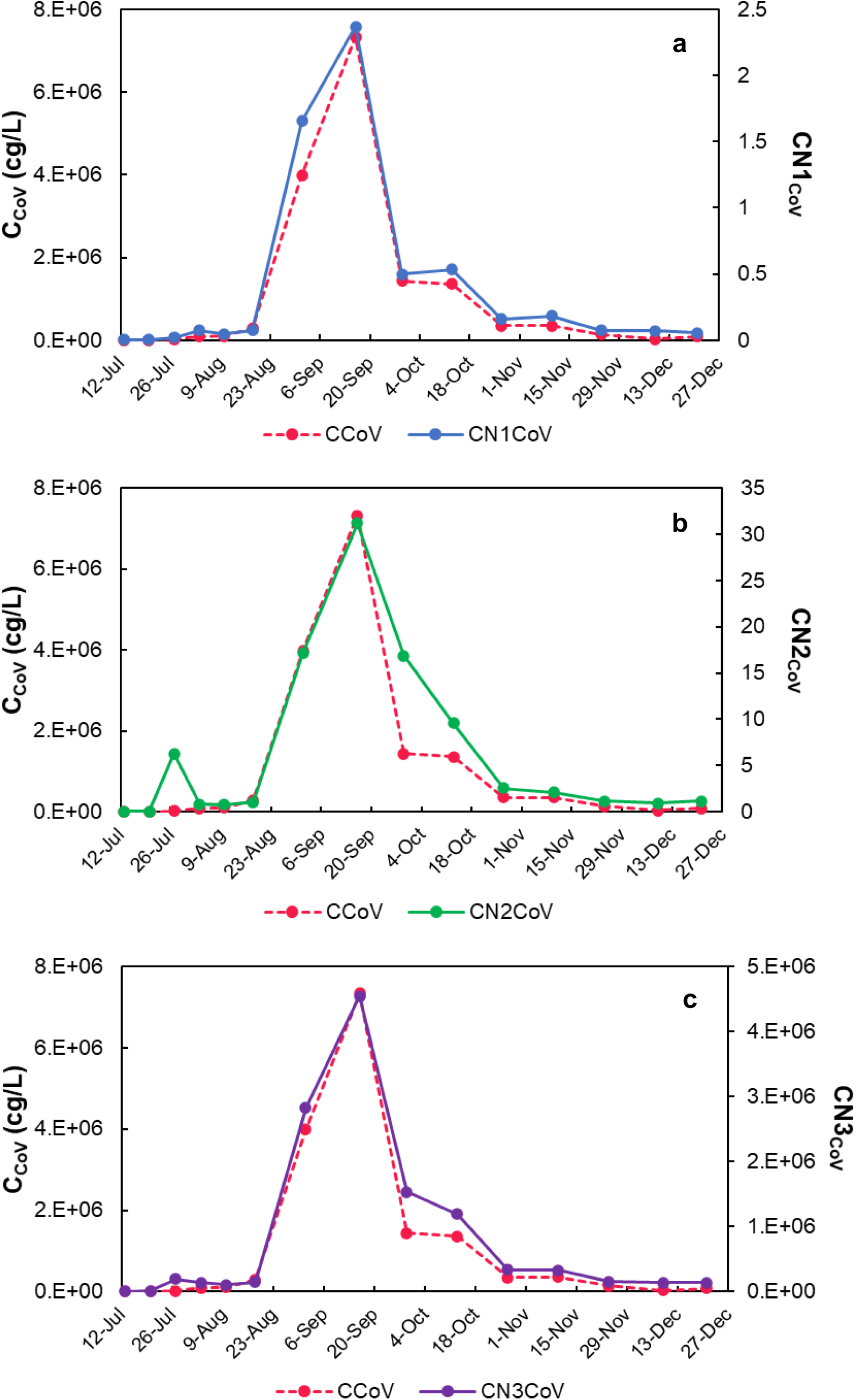
Concentration of SARS-CoV-2 experimentally determined (C_CoV_) at AR-2 in Arenales River and the normalized concentration using three different alternatives: (a) Alternative 1 (CN1_CoV_), using the concentration of human polyomavirus (HPyV), according to equation 1, (b) Alternative 2 (CN2_CoV_), using the concentration of RNase P, according to equation 2, and (c) Alternative 3 (CN3_CoV_), using the concentration of HPyV and RNase P, according to equation 3.

Regarding the three alternatives for normalization of the SARS-CoV-2 concentration evaluated, the first one was as the ratio with the concentration of human polyomavirus (equation 1), in which case the shape of the curve was not modified. However, the scale of the resulting number was too small, and the sense of the viral concentration magnitude was lost, and the units were (gc of SARS-CoV-2/gc of HPyV). The latest also happened when the ratio of SARS-CoV-2 was calculated with RNase P (equation 2), being the units (gc of SARS-CoV-2/gc of RNase P); however, the shape of the resulting curve changed a bit due to the fluctuations of RNase P concentration. Instead, the third way of performing the normalization was using the two normalizers (HPyV and RNase P), according to equation 3. In this case, the shape of the experimental SARS-CoV-2 concentration remained constant, and the normalized values kept the sense of the concentration magnitude as well as the proper units. Thus, the normalized concentration of SARS-CoV-2, obtained using both HPyV and RNase P, was selected and compared to the number of reported COVID-19 cases in the city (Figure 8). A strong positive correlation was found by Spearman test between the number of COVID-19 cases and the normalized (alternative 3) viral concentrations at AR-2 (ρ = 0.93; *p* = 0.00054).

**Figure 8.**
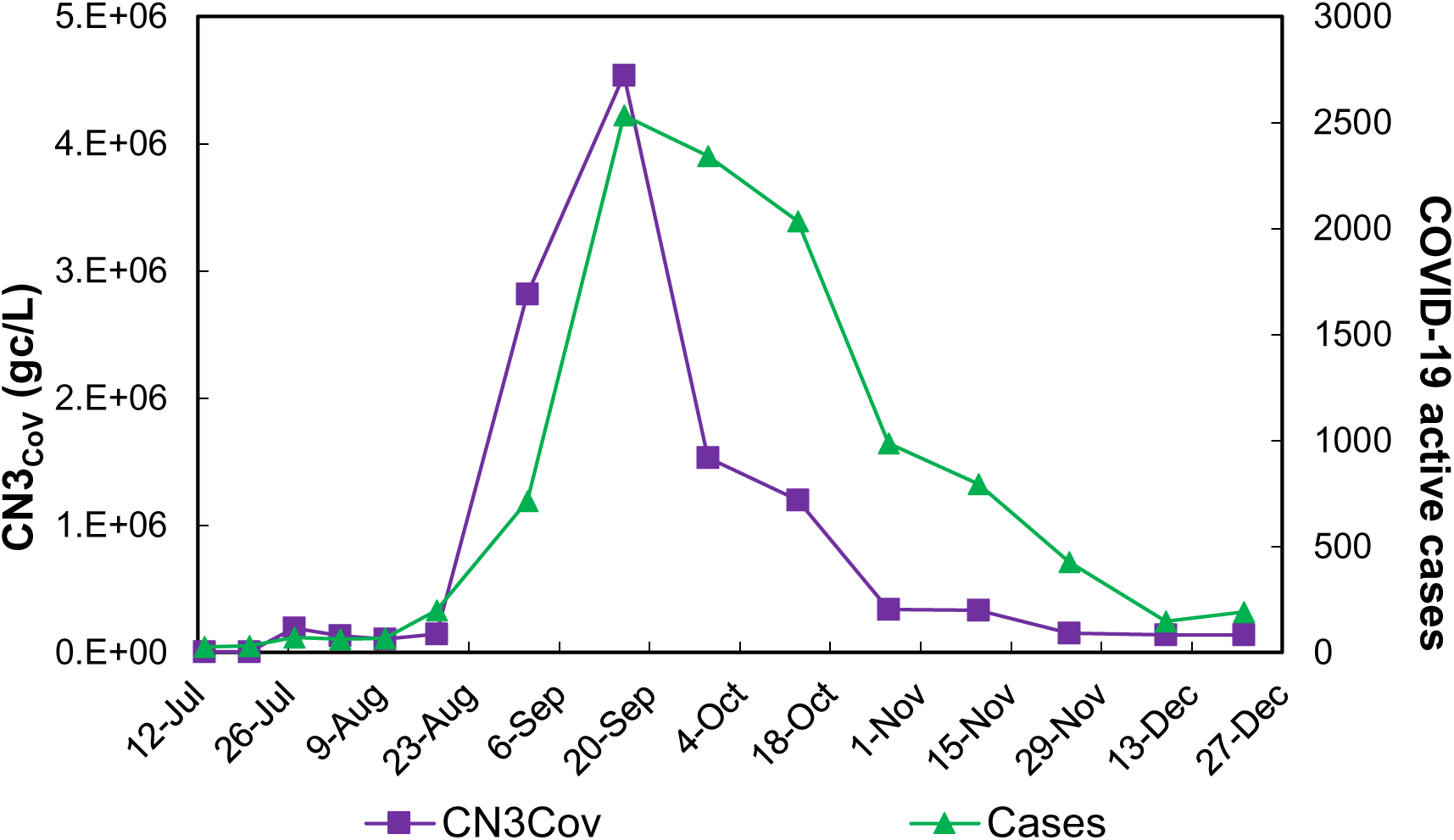
Evolution of the normalized concentration of SARS-CoV-2 (gc/L) (CN3_CoV_, according to equation 3) in the Arenales River at AR-2 and the accumulated reported COVID-19 cases, from the 12 July to 27 December 2020 during the first wave of COVID-19 in the city of Salta, Argentina.

## 4. Discussion

A dataset including experimental values for physico-chemical (four variables) and microbiological (four fecal indicator bacteria and SARS-CoV-2 concentration) variables from a total of 75 water samples from five sampling points was used to performed various bi- and multi-variable analyses. These allowed us to compare among sampling points located in the same or different rivers, to evaluate correlations, and to find the most similar and different points.

The results from the monitoring from July to December 2020, allowed to verify that from the three rivers analyzed La Caldera River has the best water quality, with low and sporadic fecal input, adequate for recreational activities, which was also described in previous work (Chávez Díaz et al, 2020; Gutiérrez Cacciabue et al, 2014). The Mojotoro and the Arenales Rivers receive the impact of the discharges of the two wastewater treatment plants of the city, WWTP-N and WWTP-S, respectively. The WWTP-N is new and has capacity for treating efficiently the flow rate from the northern part of the city, thus, the discharges do not affect the water quality of the river at least between MR-1 and MR-2. Instead, the WWTP-S is an outdated treatment plant without enough capacity for a fast-growing city. The discharges of the WWTP-S significantly impacted the Arenales River, verified at AR-2, although the water quality was already poor at AR-1, 700 m upstream, due to domestic and industrial activities and to sporadic illegal discharges of raw sewage (Poma et al., 2012). The latest was evidenced by the high concentrations of fecal bacteria indicators.

Environmental surveillance as a tool to know about the circulation of viruses and other microorganisms causing disease in the population has been proposed for decades already. In fact, rivers that were impacted by wastewater has been monitored in multiple studies, like the analyses of circulation of human polio virus as part of the effort for the eradication (Delogu et al., 2018; Iaconelli et al, 2017; WHO, 2003) and of Hepatitis A virus after vaccination campaigns (Blanco Fernandez et al, 2012a), or of evolutionary relationships between norovirus (Blanco Fernandez, 2012b).

In the case of environmental surveillance of SARS-CoV-2 almost all the efforts around the world were focused on monitoring wastewater (Medema et al., 2020; Randazzo et al., 2020b; Wu et al., 2020), most of the times analyzing the inlets (and sometimes also the outlets) of wastewater treatment plants (Gonzalez et al., 2020; Nasseri et al., 2021), but also at maintenance holes that are representative of defined areas of a city, or regions or neighborhoods (Larson et al., 2020). In some cases, the efficiency of wastewater treatment plants to remove SARS-CoV-2 was evaluated, as in the studies conducted by Wurtzer et al. (2020) in France and by Rimoldi et al. (2020) in Italy.

Wastewater samples are useful to learn about the true viral circulation in a population of geographical interest, especially to estimate the extension of virus dissemination due to asymptomatic people, that seems to play an important role in the COVID-19 pandemic (He et al., 2020). In addition, these samples present the advantages that they are anonymous and non-invasive, they do not depend on the population will (to be tested) under analysis and they can be used by the respective health authorities to make smart decisions regarding the pandemićs management (Thompson et al., 2020).

While monitoring wastewater is a useful tool to evaluate the situation in developed countries this may not be the case in threshold or developing countries, where the sewage systems are insufficient or non-existent, or when the wastewater treatment is inefficient. In most of those places, rivers or other water bodies receive the discharges of untreated (sometimes illegally) or insufficiently treated wastewater, impacting the water quality and thus, the use of the resource (Rimoldi et al., 2020). In such scenarios, surface water like natural or artificial ponds (Iglesias et al., 2020) or rivers could also be used as tools for assessing the epidemiological situation of the population. The first attempt in that direction was done by Guerrero-Latorre and coworkers (2020) that evaluated the presence of SARS-CoV-2 in a context of low sanitation countries. They sampled once in natural streams located in three different regions of Quito, Ecuador. Although they quantified SARS-CoV-2 and human adenoviruses, as well as the number of reported cases, the study is limited due to the small number of samples analyzed. Issues like the presence, persistence, and potential infectivity of SARS-CoV-2 detected in the water environment have been under the discussion for the past year (Carducci et al., 2020; La Rosa et al., 2020).

According to the results of this and previous works, the Arenales River can properly play that role as it does reflect the pathogens circulating in the population. In fact, previous studies performed in the Arenales River revealed the circulation of multiples pathogens including bacteria, parasites, and various enteric viruses (Poma et al, 2012; Blanco Fernandez et al., 2012a; Blanco Fernandez et al., 2012b; Pisano et al., 2018; Prez et al., 2020). Furthermore, the microbiological risk for the population of being in contact with those water through recreational activities was estimated for enterovirus and norovirus (Poma et al, 2019).

The representation of the viral concentration in river water allows to see the variation along time. It is like a succession of instant photos (or moments) showing the situation in that specific point under analysis. Although it allows to understand the magnitude of the viral circulation in the area, there are some limitations that should be considered, especially if the intention is to estimate the number of infected people. A water body like a river will show flow rate variations due to the input of stormwater, illegal raw sewage, and industrial effluents, among others. In other words, the viral concentration will depend on all those human or natural contributions; thus, those flow rates or dilution factors should be considered if the absolute number of virus were to be calculated. On the other hand, there could be some discussion about the influence of population dynamic in the area of impact. One possibility to account for population dynamics and for other human and non-human inputs is to normalize the viral concentration using some other target or surrogate (Medema et al, 2020b). Some chemical compounds, like fecal sterols like coprostanol (Daughton, 2012; Chen et al, 2014), other viruses common in human urinary tract, like human polyomavirus (HPyV) (McQuaig et al., 2009) or pepper mild mottle virus (PMMoV) (D’Aoust et al, 2020), or bacterial fecal indicators like human Bacteroidales (D’Aoust et al, 2020), just to mention some, have been suggested for normalization. The advantage of using any of them is that, as they are excreted by all the population all the time, then cultural or seasonal effects could be neglected.

In addition to the detection of SARS-CoV-2, two other targets linked to human contamination were quantified in this work. One of them was the gene that encodes for human RNase P, which was used in many studies as an endogenous internal control (Arnold et al., 2020; Boddicker et al., 2007; Dahdouh, et al., 2020), and it has been extensively used as a sample quality control in clinical tests in the current COVID-19 pandemic. The second one was the marker for human polyomaviruses, which has been proposed as a viral indicator of human fecal contamination and it is known to be stable in the environment. In addition, these viruses are excreted by feces and urine fluids (McQuaig et al., 2009; Bofill-Mas et al., 2000), with the species JC and BK reported as the most frequent and prevalent HPyV detected in sewage samples of this geographical region (Barrios et al., 2018).

Neither RNase P nor HPyV were detected in La Caldera River, showing again that fecal human contamination was just sporadic. In the case of Mojotoro River, RNase P was not detected and HPyV was present only in some of the samples, proving that those waters were only slightly impacted by wastewater. Conversely, in the case of the Arenales River, highly impacted by wastewater discharges, HPyV was quantified at both sampling points and the concentration was persistent and steady along the monitoring period. This was also the case for RNase P, which was only detected at AR-2.

Regarding normalization, although most of the times it has been done with only one target as reference, calculating the ratio between the gene of interest and the normalizer (McQuaig et al., 2009), it is highly recommended to use multiple genes for a more accurate normalization. For averaging of the control genes, it is recommended to use the geometric instead of the arithmetic mean, as the former controls are better for possible outlying values and abundance differences between the different genes (Vandesompele et al., 2002).

In this case the concentration of SARS-CoV-2 from AR-2 was normalized in three different ways. The first one was as the ratio with the concentration of human polyomavirus, in which case the shape of the curve was not modified, the scale of results became too small, and the sense of the viral concentration magnitude was lost, plus the units were gc of SARS-CoV-2 per gc of HPyV. A similar situation resulted when the ratio of SARS-CoV-2 was calculated with RNase P (gc of SARS-CoV-2/gc of RNase P), although the curved was slightly modified due to the fluctuations of RNase P concentration. Instead, the third way of normalizing, using both HPyV and RNase P according to equation 3, seemed more appropriate as the shape of the original concentration curved was not changed but mostly because the normalized values kept the sense of the magnitude.

In the case of the Arenales River the flow rate fluctuations were not measured; however, the difference in flow rate between the sampling points AR-1 and AR-2 (separated 700 m, no additional inputs verified) are only due to the discharges of the WWTP-S. What is interesting is that this discharge, far from producing a “dilution effect”, decreasing the viral concentration, has an “addition effect” increasing it due to the huge fecal contribution. This effect observed in the concentration of SARS-CoV-2 was also reflected in the concentration of the bacterial indicators measured. The magnitude of the discharge modified substantially, in a short distance, the microbiological and physico-chemical characteristics of the water river, differentiating this sampling point from all the others that were analyzed and minimizing any other effect related to the population dynamic or to seasonal effects. Finally, strong correlations were found between both the experimental and the normalized SARS-CoV-2 concentrations and the curve of active COVID-19 cases in the city of Salta. In this way, the water from the Arenales River, at AR-2, reflected the population situation regarding viral circulation of COVID-19 cases, therefore, it could be well used as a surveillance tool.

This is, to the best of our knowledge, the first study that showed the dynamic of SARS-CoV-2 concentration in an urban river highly impacted by wastewater and proved that it can be used for SARS-CoV-2 surveillance to support health authoritieś decisions.

## 5. Conclusions

- From the three rivers analyzed La Caldera River had the best water quality, and indicator bacteria were within acceptable limits for recreational activities.
- Although Mojotoro River receives the discharges of an old wastewater stabilization pond before MR-1 and the discharges of the northern wastewater treatment plant of the city before MR-2, they did not have a negative effect on the water quality.
- The Arenales River presented the poorest water quality. In addition to the contamination existing at AR-1 the discharges of the southern wastewater treatment plant contributed to the significant increase of fecal contamination at AR-2.
- SARS-CoV-2 was found in about half of the samples analyzed in low concentrations in La Caldera and Mojotoro Rivers. Instead, the virus concentration in the Arenales River was persistently high.
- Two human tracers, human polyomavirus (HPyV) and RNase P, were analyzed. None of them was detected in La Caldera River, only HPyV was found in Mojotoro River and at AR-1 in Arenales River, and both were quantified at AR-2 (Arenales River).
- The Arenales River, at AR-2, was highly impacted by wastewater and the concentration of SARS-CoV-2 was normalized using both human tracers. The experimental and the normalized concentrations correlated with the curve of accumulated reported COVID-19 cases in the city. Thus, the Arenales River at AR-2 reflects the epidemiological situation of the city.

## Data Availability

This is not a medical study but environmental, thus points 2, 3 and 4 are not applicable

## Acknowledgements

This research was funded by Project COVID-19 233-785, from Fondo para la Investigación Científica y Tecnológica (FONCyT), Agencia Nacional de Promoción de la Investigación, el Desarrollo Tecnológico y la Innovación, Argentina. María Noel Maidana Kulesza, Diego Gastón Sanguino-Jorquera, Sarita Reyes, María del Milagro Said-Adamo, and Martín Mainardi Remis are recipients of doctoral fellowships from CONICET.

